# Depression, anxiety and stress among elderly people living in selected old age homes in Dhaka and Gazipur, Bangladesh

**DOI:** 10.64898/2026.07.22.26358722

**Authors:** Rubaiya Binte Kabir, Md. Allama Faysal, Shamsun Nahar

**Author notes:** Corresponding author: (RBK).

## Abstract

Considering the fact of growing elderly population in Bangladesh and breaking of the cultural tradition of family taking care of the elder generation, institutionalization has become popular. This cross-sectional study was designed to evaluate the level of depression, anxiety and stress among the elderly people living in OAHs and impact of sociodemographic, anthropometric, clinical characteristics and satisfaction levels about the institutional facilities on their depression, anxiety, stress levels. Three old age homes were selected by purposive sampling-Probin Nibash (Agargaon), Priyojon Nibash (Darus Salam Road) and Old Rehabilitation Center (Gazipur). A total of 175 participants were selected from these institutions, who met inclusion criteria. They were interviewed face-to-face with a semi-structured questionnaire. DASS-21 was used to evaluate their level of depression, anxiety and stress level. SPSS version 26 was used for statistical analysis. Most prevalent age group was found to be 66-70 years with a mean age of 75.31±7.72. Female participants (52%) were more than male respondents (48%). Most of the participants was graduates (38.3%). Most of the participants (90.3%) were widowed with 1-2 children (80%). Half of the participants (50.3%) had non-government jobs previously, whereas 40% were housewives. Regarding communication, 68% never communicate with their family and most of them (80.6%) had no one to take care at their homes. More than half of the respondents (58.3%) had chronic diseases like hypertension (16.67%), diabetes (47.06%), heart disease (7.84%) etc. Majority of the participants were satisfied with the facilities like food (61.14%), recreation (83.43%), medical care (80.0%), caregiver services (96.57%), safety (98.86%), physical exercise (93.43%), social activities within the institution (99.43%) and funeral system (83.43%) provided by the institutions. Among all the participants, 80.57% had mild to extreme severe level depression, 68% had anxiety and 70.29% had stress. Depression was found to be associated significantly with female gender, less communication with family, 1-5 years of staying at OAHs (p<0.001). Anxiety was significantly associated with female gender and less communication with family (p<0.05). Stress was significantly associated with less communication with family, and 1-5 years of staying at OAHs (p<0.001).

## Introduction

Increasing life expectancy has a crucial impact on reshaping the age structure of population all over the world. In developed societies, the beginning of the old age is roughly considered around the retirement age, which is 65 years [1]. Elderly people aged 60 and above are elderly in Bangladesh. But because of poverty, physical hard working, malnutrition, geographical condition, inability and illness, people in this country become older way before the age of 60 [2]. Though only around 6% of the total population of Bangladesh constitutes the elderly population, but the rate is ever increasing.

Bangladesh is a country of long cultural and religious tradition of looking after the elderly. Family members and communities are expected to take care of their own elderly members. However, now-a-days, rapid socioeconomic and demographic transitions have been observed. As a result, changing values, the influence of western culture, and other factors have broken down the family and community care system. In this scenario the concept of Old Age Homes (OAHs) is gaining drive, and the number of people seeking institutionalization is rapidly increasing. The lack of family support makes them resort to OAHs run for their care and support. Additionally, the brochures provided by the institutions of old age homes often offer better living than living in a child’s home as an undesirable burden.

Being elderly might bring with it physiological, functional, social, cognitive, as well as financial losses. These losses make them feel environmental isolation, a subjective feeling of loneliness, anxiety, depression, and frequently, loss of motivation to continue living. On the other side, the more the rise in life expectancy and the multitude of losses, the more is the tendency to be institutionalized in old age homes in the society. Such living arrangements may have a negative impact on their mental health, because in this pattern of living, along with above mentioned losses, they also lose control over their own lives and become unable to decide regarding daily issues. In a developing country like ours, most of the elderly people suffer from some basic human problems, such as poor health and medicine facilities, exclusion and negligence, and socioeconomic insecurity [1,3]. They experience various forms of deprivations and feel a sense of worthlessness and loneliness. This leads to elderly people becoming victims of various mental problems. It is evident that hopelessness, helplessness, and depression among residents of nursing homes are significantly higher compared to those living in the community [4]. Among these disorders, depression is the most common public health problem associated with morbidity and disability among the elderly. Among older primary care patients with depression, 61.4% also have an anxiety disorder [5].

Nevertheless, the satisfaction and dissatisfaction with institutional facilities may have an impact on the mental health status among elderly living in old age homes. Yet not much is known about the response of its residents to institutionalization and its impact on their mental health causing depression, anxiety and stress [6].

Hence, the sense of wellbeing in terms of the state of mental health of the elderly people in old age homes is a thought-provoking field of inquiry. In this context, the present study was an attempt to explore the level of depression, anxiety and stress among elderly people living in old age homes in Bangladesh.

## Rationale

In the developed countries, to support the elderly people, they have systematic support services, such as, old homes, day-care centers, residential care home, nursing home etc. But in developing countries like ours, these facilities are barely available. In Bangladesh, majority of the elderly people face difficulties due to lack of sufficient income and employment opportunities, malnutrition, chronic diseases, absence of proper health care facilities and lack of adequate family support [7]. A national policy on older persons has been developed in 2013 to ensure the rights of the elderly persons where it is indicated that it is one of the fundamental duties to secure the right to enjoyment of the basic rights by the elderly people [8]. The government of Bangladesh also introduced the ‘Maintenance of Parents Act, 2013’ targeting older citizens, an important legislation for securing the basic necessities of the hopeless parents. However, evidence suggests that most of the elderly care needs remain unaddressed or overlooked [9].

Though the government, NGOs, welfare societies have come forward in these regards, these organizations only emphasize shelter, foods, cloth, physical health care that are assumed to meet all their needs. They have not sufficiently addressed mental health care services as well. As a result, those who are motivated to live in these elderly care homes do not receive enough service facility to change their life quality.

It is reported that more than half of old age home inmates suffer from one or other mental health problems, the most common being depression [10]. It is evident from the review of literature that the prevalence of psychiatric illness appears to be more in the elderly living in old age homes. But there are very few studies addressing mental health status among elderly people in OAH and associated factors causing poor status of mental health.

Thus, this study was proposed to find out the level of depression, anxiety and stress among the elderly people living in OAHs and impact of sociodemographic as well as anthropometric, clinical characteristics and satisfaction levels about the institutional facilities on their depression, anxiety and stress levels.

## Material and methods

This cross-sectional study was conducted in six months period.

### Study population and sample size calculation

The study population comprised elderly individuals residing in Probin Nibash (Agargaon) and Priyojon Nibash (Darus Salam Road) in Dhaka, as well as Old Rehabilitation Care in Gazipur. All available residents meeting the inclusion criteria (aged 60 years and above residing in the selected old homes, without diagnosed severe psychiatric problems, living there for more than a month, and were willingly participated and gave informed consent) were enrolled in the study.

Sample size was calculated using the formula n= z^2^pq/d^2^, where prevalence of depression at old age homes was considered 15.3% at 95% confidence interval [11]. the calculated sample size was 228 including 10% non-response rate. However, 175 participants were included in this study due to unavailability of the residents at the OAHs and time and resource constrains.

### Data collection instrument, method and data analysis

A semi-structured questionnaire was developed based on the objectives of the study and was used to collect data. Face-to-face interviews were conducted to fill the questionnaire. The Depression, Anxiety and Stress Scale - 21 Items (DASS-21) was used to measure the emotional states of depression, anxiety and stress. Coding was done by giving a serial number for each answer.

Data were analyzed using the Statistical Package for the Social Sciences (SPSS), version 27. Descriptive statistics were calculated using frequency distribution, including mean, standard deviation, and percentage. Chi-square tests with cross-tabulation were performed to assess the association between independent variables (demographic characteristics) and outcome variables, with a 95% confidence interval. A p-value less than 0.05 was considered statistically significant. Results were presented using frequency tables, graphs, and charts.

### Ethical issues

Approval from the National Research Ethics Committee of the Bangladesh Medical Research Council, Bangladesh was obtained (BMRC/Revenue/Research Grant/2026/646(01-168). Permission was taken from the authorities of selected organizations. Written informed consent was obtained from the respondents prior to data collection. The objectives of the study were briefly explained to them before consent was taken. Privacy and confidentiality were strictly maintained. Respondents had the right to withdraw from the study at any time during data collection.

## Results

Table 1 demonstrates sociodemographic distribution of participants. Most prevalent age group was 66-70 years (26.3%), followed by 24.6% grouped in >80 years age group. Their mean age was 75.31 ± 7.72. Females were slightly more in number than the male participants, 52 and 48% respectively. Most of the participants was graduates (38.3%) followed by illiterate (22.3%). most of the participants (90.3%) were widowed and 80% had 1-2 children; among those who had children (83.4%), most of them had only female children (59.18%). Half of the participants (50.3%) had non-government job previously. Among all the participants, 83.4% were supported by the institution, all of whom lived in Old Rehabilitation Care. Nine (5.1%) of the participants were supported by their children, all of them lived in Priyojon Nibash. Participants who were living in Probin Nibash either were self-employed (72.7%) or depends on pension (27.3%).

**Table 1:** Socio-demographic distribution of participants (N= 175):

|  | Frequency | Percentage (%) |
| --- | --- | --- |
| Age distribution (N= 175) |  |  |
| 60- 65 years | 15 | 8.6 |
| 66- 70 years | 46 | 26.3 |
| 71- 75 years | 30 | 17.7 |
| 76- 80 years | 40 | 22.9 |
| >80 years | 43 | 24.6 |
| Mean ± SD | 75.31 ± 7.72 |  |
| Gender distribution (N= 175) |  |  |
| Male | 84 | 48 |
| Female | 91 | 52 |
| Highest education level (N= 175) |  |  |
| Nonformal/ only read & write | 16 | 9.1 |
| Illiterate | 39 | 22.3 |
| SSC | 18 | 10.3 |
| HSC | 17 | 9.7 |
| Graduation | 67 | 38.3 |
| Masters | 17 | 9.7 |
| 6-9 | 1 | .6 |
| **Marital status (N= 175)** |  |  |
| Never married | 6 | 3.4 |
| Married | 3 | 1.7 |
| Widowed | 158 | 90.3 |
| Refused | 8 | 4.6 |
| **Number of children of the participants (N= 175)** |  |  |
| Nil | 28 | 16 |
| 1-2 | 141 | 80 |
| >2 | 6 | 3.4 |
| **Details of the children (n=147)** |  |  |
| Male | 40 | 27.21 |
| Female | 87 | 59.18 |
| Both | 20 | 13.61 |
| **Previous occupation (N- 175)** |  |  |
| Govt job | 10 | 5.7 |
| Non govt job | 88 | 50.3 |
| Day labor | 4 | 2.3 |
| Business | 3 | 1.7 |
| Housewife | 70 | 40.0 |
| **Resident old age homes (N= 175)** |  |  |
| Old Rehabilitation Care | 146 | 83.4 |
| Priyojon Nibash | 18 | 10.3 |
| Probin Nibash | 11 | 6.3 |

| **Source of income (N= 175)** |  |  |
| --- | --- | --- |
| Institution | 146 | 83.4 |
| Children | 9 | 5.1 |
| Self- employed | 11 | 6.3 |
| Pension | 4 | 2.3 |
| Other Relatives | 5 | 2.9 |

Table 2 shows the frequency of communication of the participants with the families. Among all the participants, 68% of their families never communicate with them, which is a substantial proportion.

**Table 2:** Distribution of respondents by frequency of communication with their family (n=175):

| **Frequency of communication** | **Frequency (n)** | **Percentage (%)** |
| --- | --- | --- |
| **Never** | 119 | 68.0 |
| **Rarely** | 24 | 13.7 |
| **Sometimes** | 23 | 13.1 |
| **Frequently** | 9 | 5.1 |
| **Total** | **175** | **100.0** |

About 81% admitted that they have no one to take care of them at home for which they chose to live at old age homes which is described in Table 3.

**Table 3:** Distribution of respondents by reasons for living in old homes (n=175)

| **Reasons** | **Frequency (n)** | **Percentage (%)** |
| --- | --- | --- |
| **Negligence of family members** | 16 | 9.1 |
| **No care taking person at home** | 141 | 80.6 |
| **Health problem** | 5 | 2.9 |
| **Death of spouse** | 1 | .6 |
| **Personal freedom** | 12 | 6.9 |
| **Total** | **175** | **100.0** |

The most of the participants (67.4%) were living less than 5 years (Table 4).

**Table 4:** Distribution of participants according to duration of stay at the old age homes (n=175)

| **Duration of stay** | **Frequency (n)** | **Percentage (%)** |
| --- | --- | --- |
| Less than 5 years | 118 | 67.4 |
| 5- 10 years | 24 | 13.7 |
| 10- 15 years | 19 | 10.9 |
| 16- 20 years | 13 | 7.4 |
| More than 20 years | 1 | .6 |
| **Total** | **175** | **100.0** |

Table 5 demonstrates the distribution of chronic diseases among the participants. This table shows that 102 (58.3%) participants had chronic diseases.

**Table 5:** Distribution of participants by presence of chronic disease (n=175):

| **Presence of disease** | **Frequency (n)** | **Percentage (%)** |
| --- | --- | --- |
| Yes | 102 | 58.3 |
| No | 73 | 41.7 |
| **Total** | **175** | **100.0** |

Among those who had chronic diseases, 47.06% had diabetes, followed by 16.67% HTN, and then other chronic diseases (Table 6).

**Table 6:** Distribution of participants by their chronic disease (n=102):

| **Types of diseases** | **Frequency (n)** | **Percentage (n)** |
| --- | --- | --- |
| Diabetes | 48 | 47.06 |
| HTN | 17 | 16.67 |
| Diabetes + HTN | 18 | 17.65 |
| Heart disease | 8 | 7.84 |
| COPD | 1 | 0.98 |
| Eyesight problem | 3 | 2.94 |
| Others | 7 | 6.86 |
| **Total** | **102** | **100.0** |

Majority of the participants (62.9%) had normal nutritional status (Table 7).

**Table 7:** Distribution of participants by their nutritional status (n= 175):

| **Nutritional status** | **Frequency (n)** | **Percentage (%)** |
| --- | --- | --- |
| Normal | 110 | 62.9 |
| Overweight | 55 | 31.4 |
| Obesity | 10 | 5.7 |
| **Total** | **175** | **100.0** |

Table 8 shows the satisfaction levels about different facilities of the institutions among the participants. Most of the participants (61.14-99.43%) were satisfied with the overall facilities of the institutions.

**Table 8:** Satisfaction level about different facilities of the institutions (n=175):

|  | **Satisfied**<br>**n (%)** | **Average**<br>**n (%)** | **Dissatisfied**<br>**n (%)** | **Neither**<br>**satisfied nor**<br>**dissatisfied**<br>**n (%)** | **Total**<br>**n (%)** |
| --- | --- | --- | --- | --- | --- |
| **Food** | 107 (61.14) | 59 (33.71) | 6 (3.4) | 3 (1.71) | 175 (100) |
| **Recreational**<br>**facilities** | 146 (83.43) | 28 (16.0) | 0 | 1 (0.57) | 175 (100) |
| **Medical Care** | 140 (80.0) | 28 (16.0) | 6 (3.4) | 1 (0.57) | 175 (100) |
| **Service of the**<br>**caregivers** | 169 (96.57) | 5 (2.86) | 0 | 1 (0.57) | 175 (100) |
| **Safety** | 173 (98.86) | 2 (1.14) | 0 | 0 | 175 (100) |
| **Facilities of**<br>**physical exercise** | 163 (93.14) | 10 (5.71) | 0 | 2 (1.14) | 175 (100) |
| **Involvement with**<br>**social activities** | 174 (99.43) | 1 (0.57) | 0 | 0 | 175 (100) |
| **Funeral facilities** | 146 (83.43) | 0 | 0 | 29 (16.57) | 175 (100) |

Figs 1, 2 and 3 demonstrates the proportion of depression, anxiety and stress among elderly participants depending on the DASS-21 respectively. Among all the participants, 80.57% had mild to extremely severe level of depression, most being 42.29% extremely severe (Fig 1). Out of 175 participants, 68% had anxiety (Fig 2). Among all the participants, 70.29% had stress of various level (Fig 3).

**Fig 1:**
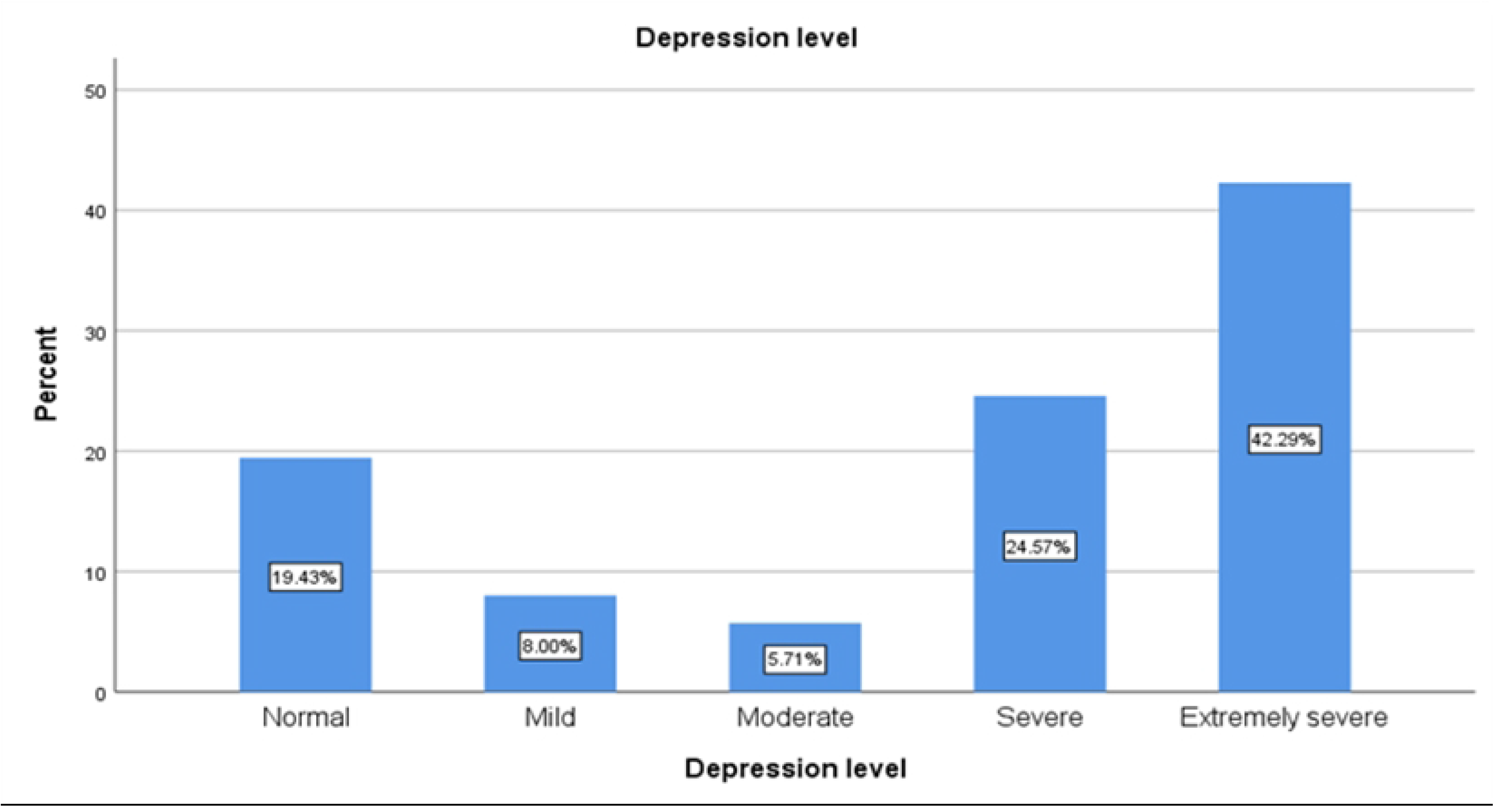
Frequency of depression among the participants (n=175)

**Fig 2:**
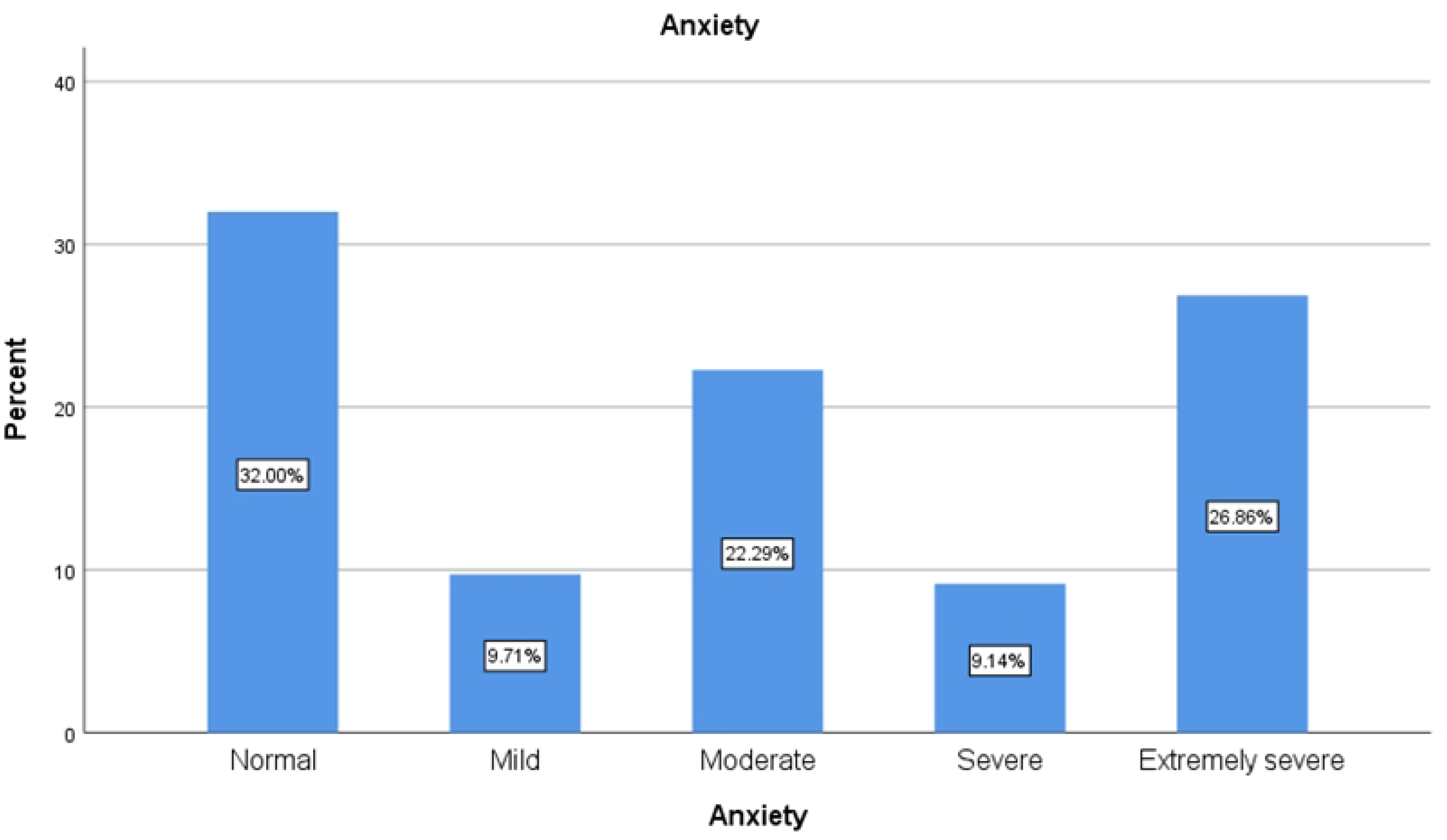
Frequency of anxiety among the participants (n=175)

**Fig 3:**
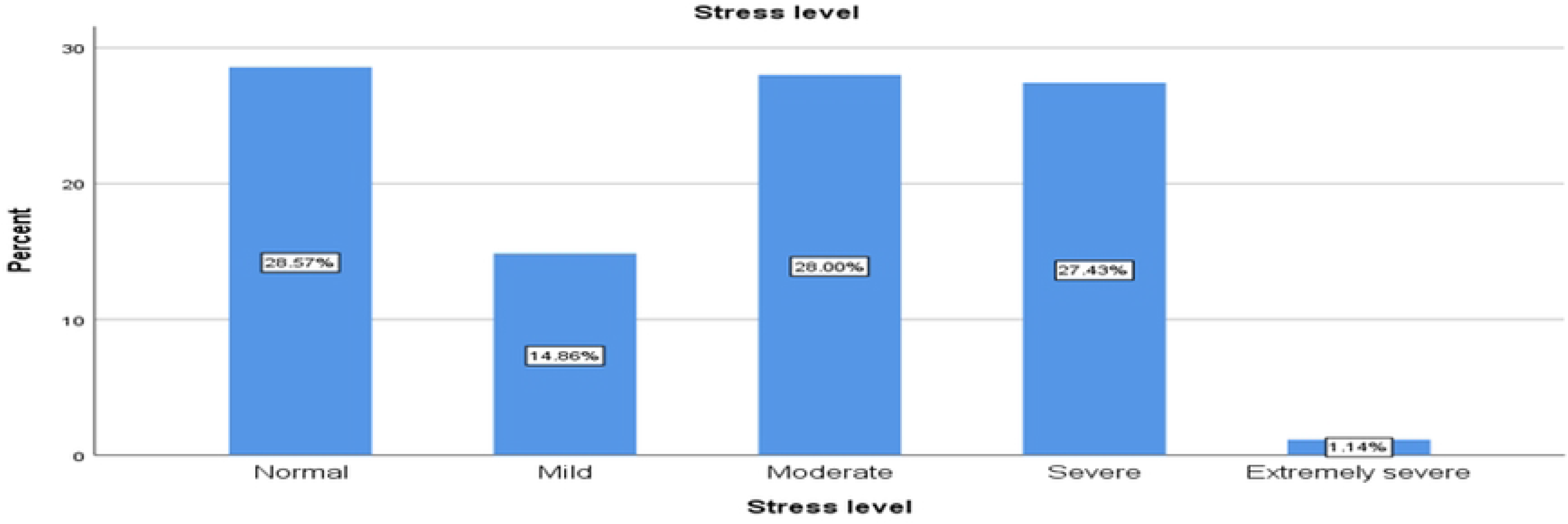
Frequency of stress among the participants (n=175)

Table 9, 10 and 11 demonstrate the bivariate analysis of different factors with depression, anxiety and stress respectively. In table 9, age was not significantly associated with the outcome. However, gender, communication with family, duration of staying at OAH, and chronic disease were significantly associated with depression. Male participants were less likely to have depression compared to females (OR = 0.18, 95% CI = 0.07–0.43, p < 0.001). Participants who communicate rarely (OR = 8.05, p = 0.004) and sometimes (OR = 6.25, p = 0.03) with their families are prone to develop depression which is statistically significant compared to those who frequently communicate. Duration of staying at OAH showed significant association with depression, though unstable odds ratios was observed due to small sample sizes and zero cell counts. Participants who did not have chronic disease were significantly associated with depression (OR = 0.19, 95% CI = 0.07–0.51, p = 0.001).

**Table 9:** Bivariate association between different factors with depression:

|  | No | Yes | Sig | OR | Lower | Upper |
| --- | --- | --- | --- | --- | --- | --- |
| **Age** |  |  |  |  |  |  |
| 66- 70 years | 7 (15.2) | 39 (84.8) | .75 | .69 | .07 | 6.78 |
| 71- 75 years | 4 (12.9) | 27 (87.1) | .75 | .74 | .13 | 4.44 |
| 76- 80 years | 3 (7.5) | 37 (92.5) | .14 | 4.45 | .62 | 32.01 |
| >80 years | 15 (34.9) | 28 (65.1) | .55 | 1.79 | .26 | 12.35 |

| **Gender** |  |  |  |  |  |  |
| --- | --- | --- | --- | --- | --- | --- |
| Female | 7 (7.7) | 84 (92.3) |  |  |  |  |
| Male | 27 (32.1) | 57 (67.9) | .000 | .17 | .072 | .43 |
| **Communication with family** |  |  |  |  |  |  |
| Never | 16 (13.4) | 103 (86.6) | .003 |  |  |  |
| Rarely | 4 (16.7) | 20 (83.3) | .004 | 8.05 | 1.952 | 33.17 |
| Sometimes | 9 (39.1) | 14 (60.9) | .03 | 6.25 | 1.145 | 34.12 |
| Frequently | 5 (55.6) | 4 (44.4) | .40 | 1.94 | .409 | 9.24 |
| **Duration of staying at OAH** |  |  |  |  |  |  |
| 1-5 years | 13 (11.0) | 105 (89.0) | .000 |  |  |  |
| 6-10 years | 4 (16.7) | 20 (83.3) | 1.000 | 13047763337.71 | .000 | . |
| 11- 15 years | 10 (52.6) | 9 (47.4) | 1.000 | 8077186828.106 | .000 | . |
| 16- 20 years | 6 (46.2) | 7 (53.8) | 1.000 | 1453893629.059 | .000 | . |
| >20 years | 1 (100) | 0 | 1.000 | 1884676926.558 | .000 | . |
| **Chronic disease** |  |  |  |  |  |  |
| Absence | 5 (6.8) | 68 (93.2) |  |  |  |  |
| Presence | 29 (28.4) | 73 (71.6) | .001 | .185 | .068 | .51 |

**Table 10:** Bivariate association between different factors with anxiety:

|  | No | Yes | Sig. | OR | Lower | Upper |
| --- | --- | --- | --- | --- | --- | --- |
| **Age** |  |  |  |  |  |  |
| 60-65 | 6 (40.0) | 9 (60.0) | .16 |  |  |  |
| 66-70 | 11 (23.9) | 35 (76.1) | .61 | 0.65 | 0.12 | 3.48 |
| 71-75 | 15 (48.4) | 16 (51.6) | .77 | 1.19 | 0.37 | 3.84 |
| 76-80 | 5 (12.5) | 35 (87.5) | .22 | 0.44 | 0.11 | 1.66 |
| >80 | 19 (44.2) | 24 (55.8) | .13 | 3.07 | 0.71 | 13.26 |
| **Gender** |  |  |  |  |  |  |
| Female | 10 (11.0) | 81 (89.0) |  |  |  |  |
| Male | 46 (54.8) | 38 (45.2) | .000 | 0.048 | 0.017 | 0.139 |
| **Communication with the family** |  |  |  |  |  |  |
| Never | 33 (27.7) | 86 (72.3) | .14 |  |  |  |
| Rarely | 6 (25.0) | 18 (75.0) | .07 | 6.23 | 0.89 | 43.87 |
| Sometimes | 12 (52.2) | 11 (47.8) | .31 | 3.35 | 0.32 | 34.80 |
| Frequently | 5 (55.6) | 4 (44.4) | .48 | 2.08 | 0.27 | 15.94 |
| **Duration of staying in OAH** |  |  |  |  |  |  |
| 1-5 years | 32 (27.1) | 86 (72.9) | .099 |  |  |  |
| 6-10 years | 6 (25.0) | 18 (75.0) | 1.00 | 10464458244.07 |  |  |
| 11-15 years | 10 (52.6) | 9 (47.4) | 1.00 | 8407006161.82 |  |  |
| 16-20 years | 7 (53.8) | 6 (46.2) | 1.00 | 1946052342.81 |  |  |
| >20 years | 1 (100) | 0 | 1.00 | 1326971000.59 |  |  |
| **Chronic disease** |  |  |  |  |  |  |
| Absence | 18 (24.7) | 55 (75.3) |  |  |  |  |
| Presence | 38 (37.3) | 64 (62.7) | .095 | 0.41 | 0.15 | 1.17 |

**Table 11:**
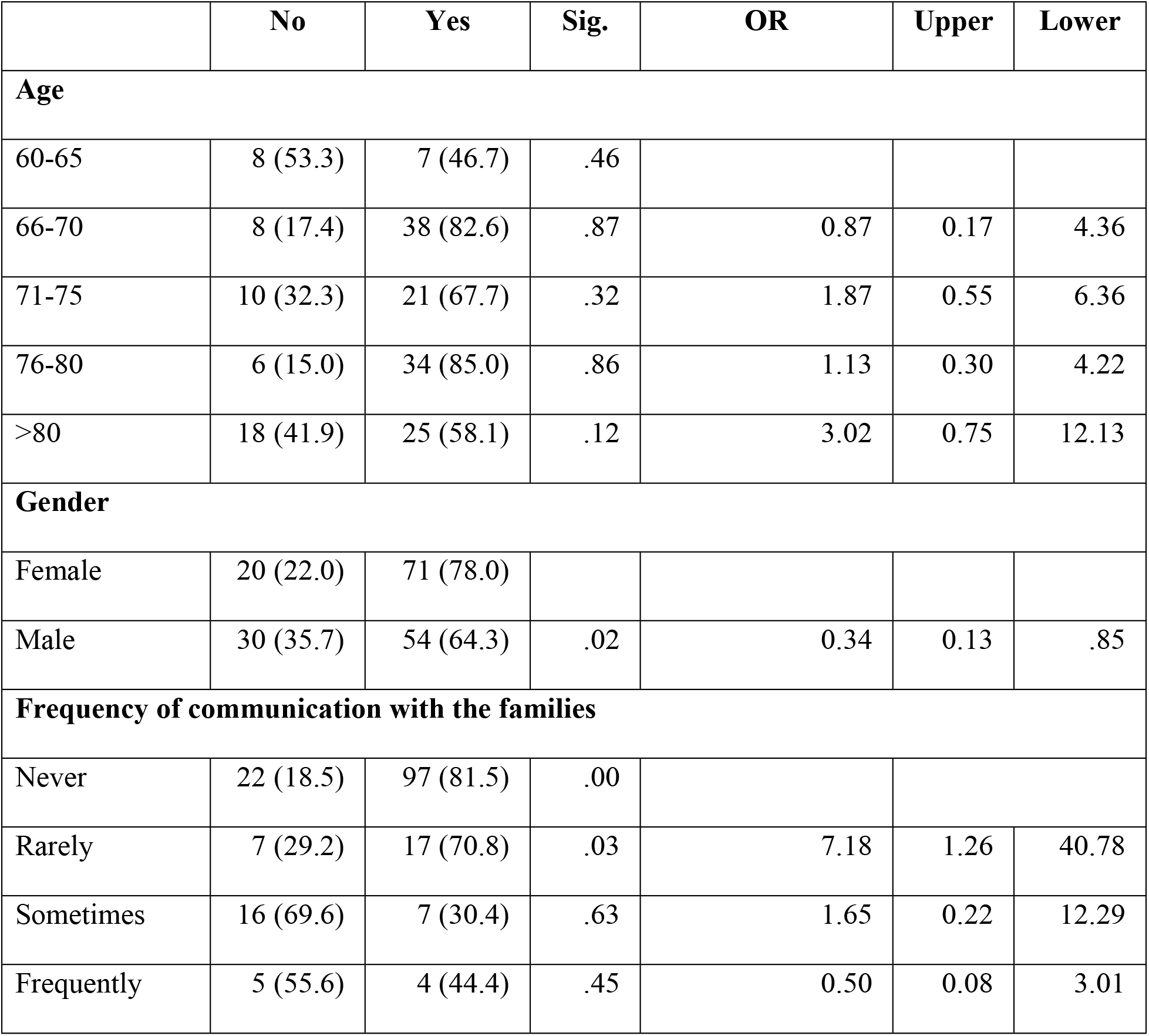
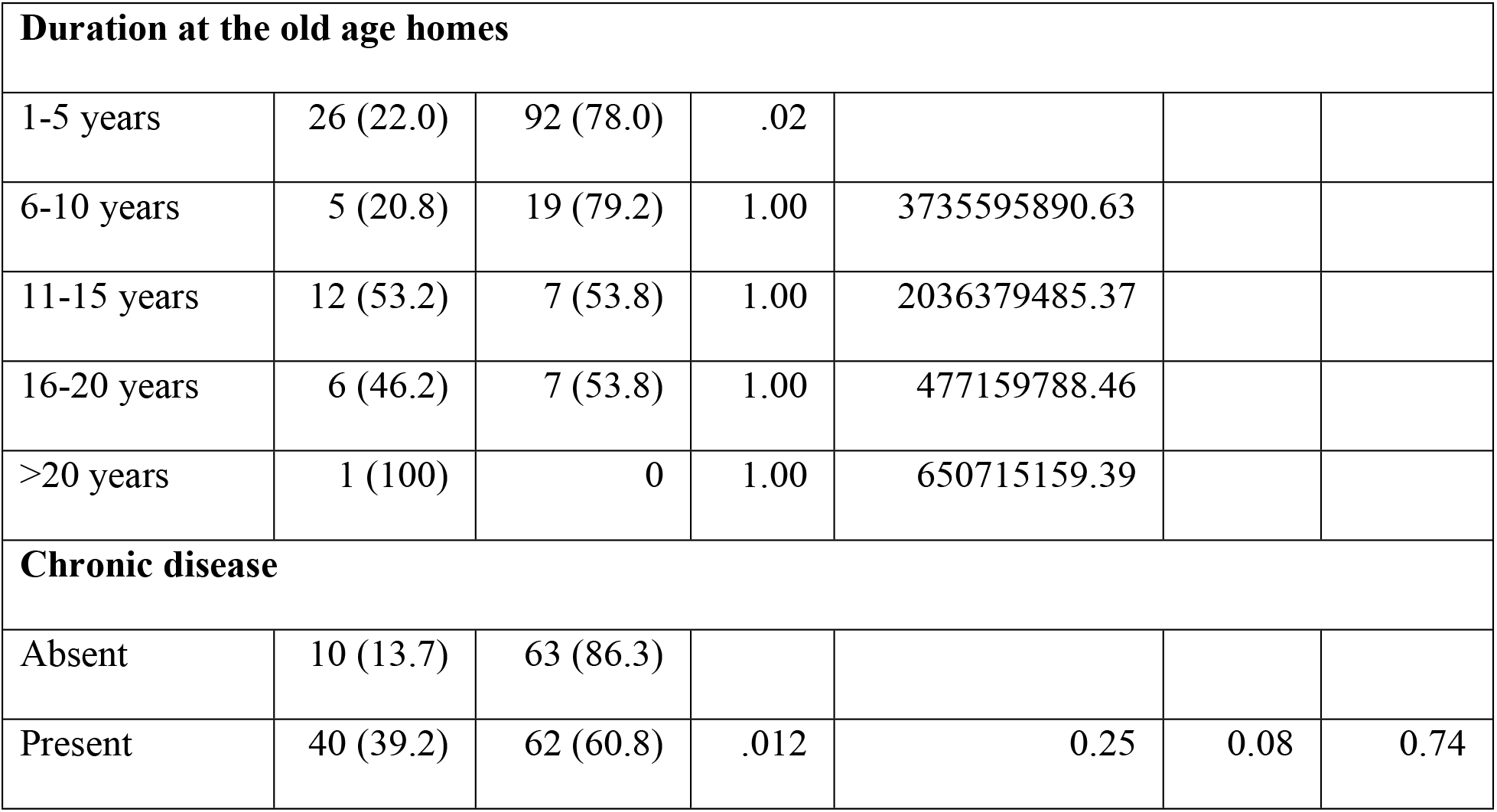
Bivariate association between different factors with stress:

|  | No | Yes | Sig. | OR | Upper | Lower |
| --- | --- | --- | --- | --- | --- | --- |
| **Age** |  |  |  |  |  |  |
| 60-65 | 8 (53.3) | 7 (46.7) | .46 |  |  |  |
| 66-70 | 8 (17.4) | 38 (82.6) | .87 | 0.87 | 0.17 | 4.36 |
| 71-75 | 10 (32.3) | 21 (67.7) | .32 | 1.87 | 0.55 | 6.36 |
| 76-80 | 6 (15.0) | 34 (85.0) | .86 | 1.13 | 0.30 | 4.22 |
| >80 | 18 (41.9) | 25 (58.1) | .12 | 3.02 | 0.75 | 12.13 |
| **Gender** |  |  |  |  |  |  |
| Female | 20 (22.0) | 71 (78.0) |  |  |  |  |
| Male | 30 (35.7) | 54 (64.3) | .02 | 0.34 | 0.13 | .85 |
| **Frequency of communication with the families** |  |  |  |  |  |  |
| Never | 22 (18.5) | 97 (81.5) | .00 |  |  |  |
| Rarely | 7 (29.2) | 17 (70.8) | .03 | 7.18 | 1.26 | 40.78 |
| Sometimes | 16 (69.6) | 7 (30.4) | .63 | 1.65 | 0.22 | 12.29 |
| Frequently | 5 (55.6) | 4 (44.4) | .45 | 0.50 | 0.08 | 3.01 |

| Duration at the old age homes |  |  |  |  |  |  |
| --- | --- | --- | --- | --- | --- | --- |
| 1-5 years | 26 (22.0) | 92 (78.0) | .02 |  |  |  |
| 6-10 years | 5 (20.8) | 19 (79.2) | 1.00 | 3735595890.63 |  |  |
| 11-15 years | 12 (53.2) | 7 (53.8) | 1.00 | 2036379485.37 |  |  |
| 16-20 years | 6 (46.2) | 7 (53.8) | 1.00 | 477159788.46 |  |  |
| >20 years | 1 (100) | 0 | 1.00 | 650715159.39 |  |  |
| Chronic disease |  |  |  |  |  |  |
| Absent | 10 (13.7) | 63 (86.3) |  |  |  |  |
| Present | 40 (39.2) | 62 (60.8) | .012 | 0.25 | 0.08 | 0.74 |

Table 10 demonstrates that, there were no statistically significant association between age, communication with family, duration of staying in OAH, and chronic disease with anxiety (p > 0.05), although higher odds were observed among participants aged above 80 years and those with rare family communication. However, male participants had less anxiety compared to female participants which was statistically significant (OR = 0.05, 95% CI = 0.02–0.14, p < 0.001).

Table 11 shows, age had no significant association with the stress. Male participants were significantly less stressed than females (OR = 0.34, 95% CI = 0.13–0.85, p = 0.02). Elderly who rarely communicated with their families were more stressed (OR = 7.18, p = 0.03). Interestingly, participants with chronic disease were less stressed, which was statistically significant (OR = 0.25, 95% CI = 0.08–0.74, p = 0.01).

## Discussion

Considering the ever-increasing number of elderly people in Bangladesh, it is of utmost importance to address different aspects of mental health in addition to the physical health care. In the current study, evaluation of depression, anxiety and stress have been evaluated among the elderly people living in old age homes.

The most prevalent age group in this study was 66-70 years (26.3%) with a mean age of 75.31 ±7.72 which coincide with the study reported by Harun et al [11]. This trend is may be due to the fact that most of the residents come to OAHs at their sixties. Regarding gender distribution, this study found female participants slightly more than male participants unlike the above-mentioned study. Most of the participants were widowed. This may contribute to be the reason of staying at old care for the females, as most of the females with lower educational level completely depends on the spouses. A large portion of participants had 1-2 children and, most of them being female (59.18%). Moreover, most of the female participants were housewives (40.0%). It is assumed that, in lower socio-economic countries like Bangladesh, female married children are unable to provide for their parents due to traditional patriarchal norms. This might contribute to the reasons of keeping older parents at OAHs. It is matter of despair that, most of the participants (68%) never communicate with their family and living in the OAHs due to having no one to take care at their homes (80.6%).

Out of 175 participants, 58.3% had chronic diseases like diabetes, hypertension, heart disease, chronic pulmonary disease etc. which is a common scenario of aging process. For a better alternative of living independently not to be burden in children’s home, elderly people look forward to OAHs for facilities that can provide them financial and social security [12]. In this study, satisfaction level was assayed regarding different facilities experienced by the participants. More than half of the participants were found satisfied with the foods served from the institutions (61.14%). However, some stated the satisfaction level as average (33.71%). Residents of Probin Nibash mostly provide food for their own and only depends on the institution’s cafeteria if needed. Almost all the participants were satisfied with the safety and social activities inside the institution. Funeral facilities were available at Old Rehabilitation Center at its own graveyard which give them a sense of security and satisfaction at their final times.

Depression and anxiety are commonly psychiatric problems among the elderly people. Co-occurrence of depression and anxiety is also associated with emotional distress [13,14]. In this study substantial proportion of participants had mild to extremely severe depression (80.57%), anxiety (68%) and stress (70.29%). The prevalence does not match with the prevalence reported by Raeisvandi et al [15]. The different prevalence rate may be due to cultural, environmental, hereditary factors and study methodology [15].

Various factors may be attributed to these psychiatric conditions. Age was found to be significantly associated with depression among elderly >80 years [16]. In contrast, in this study though depression, anxiety and stress were observed to be prevalent among 76-80 years age group, the association was found to be non-significant. Different study tool, study population, study settings may be the factors behind different findings [16]. Results showed significant association of female gender with presence of depression, anxiety. However, though stress was found slightly greater among female participants, it was not statistically significant. Mali et al., reported opposing findings regarding sex [16].

In this study, the association of never or rarely communication with the family was statistically significant with depression, anxiety as well as stress. This phenomenon was also observed significantly at first 1-10 years of staying at OAHs. Residential unfamiliar environment at OAHs, living away from children, grandchildren and relatives vital cause of distress [17].

Presence of chronic disease has been reported as one of key pivotal factors affecting depression [16]. However, this study observed a contrasting result. Contradictory results may be due to different diagnostic tool, study population or study settings [18]. In current study, majority of the respondents were satisfied with different institutional facilities irrespective of the payment methods. So, it is assumed that, the depression, anxiety or stress present in respondents are not related to the institutional facilities in this particular study.

## Conclusion

Based on the results evaluated in this study, it can be concluded that, post retirement age group, widowed females who used to be housewives with low educational level were potential clients of OAHs. Residents who had female children, males who had non-government jobs were majority in number in these facilities. Depression, anxiety and stress among the elderly people residing at OAHs depend on multiple factors, where female gender, less frequent communication with family members, duration of stay, to be strongly associated with depression, anxiety as well as stress.

## Data Availability

The data set has been uploaded as an attached file under supporting information (file name: SPSS data)

## Contributorship

RBK contributed in research concept, study design, protocol writing, defending Ethical Review Committee, developing data collection tool, data collection, data analysis and interpretation, writing manuscript

MAF contributed in data analysis, reviewing manuscript

SN contributed in reviewing manuscript

## Acknowledgement

The authors would like to acknowledge the old age homes-Probin Nibash, Priyojon Nibash of Dhaka and Old Rehabilitation Center of Gazipur and the participants for permitting them to collect data. The authors are grateful for the funding received from Bangladesh Medical Research Council, Bangladesh to conduct this study.

